# REVIVE-HF: Rehabilitation with Immersive Virtual Reality and Exercise in Hospitalized Patients with Heart Failure - A Randomized Controlled Trial Protocol

**DOI:** 10.64898/2025.12.11.25342100

**Authors:** Ariele dos Santos Costa, Caroline Bublitz Barbosa, Solange Guizilini, Isis Begot Krainer, Pedro Ivo de Marqui Moraes, Vagner Rogério dos Santos, Rita Simone Lopes Moreira

## Abstract

**Background:** Heart failure (HF) is a chronic condition characterized by significant functional limitations, with exercise intolerance as a major determinant of reduced quality of life. Supervised aerobic exercise is a core intervention in cardiac rehabilitation, yet adherence and tolerance may be hindered by physical and motivational factors. Immersive technologies such as virtual reality (VR) have shown potential to increase engagement and positive perceptions during exercise. This study aims to evaluate the influence of VR on tolerance to cycle-ergometer aerobic exercise in hospitalized patients with HF.

**Methods:** We will conduct a randomized, controlled, parallel-group trial with 1:1 allocation. Adult inpatients with a diagnosis of HF will be screened for eligibility and randomly assigned to two groups: control (cycle-ergometer aerobic exercise) and intervention (the same exercise combined with an Meta Quest 2 VR device). Each participant will complete a single session of up to 20 minutes, consisting of five blocks of three minutes of continuous pedaling interspersed with one-minute passive rests, with early termination in case of intolerance. The primary outcome is effective exercise time (minutes) until interruption due to physical limitation or symptoms. Secondary outcomes include rating of perceived exertion, hemodynamic parameters, exercise enjoyment (PACES), and usability of the technology (SUS, intervention group only).

**Discussion:** This study will investigate whether VR can improve tolerance to aerobic exercise during hospitalization for HF, potentially providing preliminary evidence to guide the use of immersive technologies in cardiac rehabilitation. If benefits are observed, VR could be incorporated as a complementary tool to optimize adherence, motivation, and safety in supervised exercise programs in this clinical context.

**Trial registration:** Brazilian Clinical Trials Registry (ReBEC): RBR-4hrmkzz.

**Strengths and limitations:**

- Single-centre randomized controlled trial with concealed allocation and blinded analysis, enhancing internal validity.
- Highly standardised intervention and control conditions differing only by the presence of immersive VR, isolating the specific effect of VR.
- Single-session, single-centre design with a modest sample size limits generalisability and precludes assessment of long-term clinical outcomes.

## Introduction

### Background and rationale {9a}

Among chronic noncommunicable diseases, cardiovascular diseases (CVD) are a major public health concern (1,2), with heart failure (HF) being one of their most relevant chronic manifestations. HF is a clinical syndrome characterized by classic symptoms such as fatigue and dyspnea, which may be accompanied by signs such as jugular venous distension, pulmonary congestion, and peripheral edema. These symptoms result from structural and/or functional cardiac changes leading to reduced cardiac output and/or increased intracardiac pressures at rest or during exertion(3). Even when stable and well-compensated, HF commonly presents with exercise intolerance as a primary symptom, negatively affecting functional capacity and independence in activities of daily living, thereby impairing quality of life. Additionally, HF treatment involves complex regimens, including multiple medications, fluid control, and frequent medical visits, which may increase burden on patients and families, limit social interactions, and lead to isolation (4).

Cardiac rehabilitation (CR) is an essential intervention to improve physical, psychological, and social conditions in individuals with CVD, preserving and promoting quality of life and reducing risk factors. It is recognized as an effective and low-cost nonpharmacological treatment for patients with HF. Despite well-documented benefits and strong guideline recommendations, fewer than 20% of patients with HF participate in CR programs globally (5,6), with rates as low as 1–3% reported in Brazil (7,8). Barriers to CR participation are multifactorial and include patient-level factors (lack of knowledge, exercise fear, low self-efficacy, comorbidities, lack of motivation), logistical barriers (distance, transportation2, time conflicts, cost), health system factors (limited program availability, lack of physician referral), and environmental factors (lack of social support)(9,10)Notably, many patients perceive conventional CR programs as repetitive and monotonous, contributing to poor adherence and high dropout rates. This underscores the urgent need for innovative strategies to enhance patient engagement - particularly by promoting positive exercise experiences and exploring methods that increase motivation and facilitate adherence to physical activity, especially in resource-limited acute care settings.

Virtual reality (VR) has emerged as a promising technology to enhance CR engagement and adherence. Unlike conventional CR methods, which may be repetitive and less engaging, VR systems enable enjoyable activities with therapeutic purpose and interaction with virtual environments tailored to rehabilitation needs (11,12). VR is hypothesized to enhance engagement and adherence through several interconnected psychological and physiological mechanisms: (1) immersive environments reduce perceived exertion and distract from unpleasant physical sensations(13); (2) VR fulfills core psychological needs (autonomy, competence, relatedness) as described by Self-Determination Theory(14) ; (3) gamified elements and real-time feedback activate brain reward systems, promoting self-efficacy and behavioral persistence(15) ; and (4) immersive experiences significantly reduce anxiety while improving mood states, which may enhance willingness to participate in rehabilitation (12,16). These mechanisms suggest that VR may be particularly valuable for acutely hospitalized HF patients, who often experience high exercise-related anxiety, low motivation, and multiple barriers to traditional rehabilitation.

Preliminary evidence from our pilot study(17) demonstrated the feasibility of immersive VR exercise in hospitalized HF patients, with participants reporting enjoyment (Physical Activity Enjoyment Scale scores). However, rigorous evidence from a randomized controlled trial is needed to establish definitive clinical efficacy and justify broader implementation.

This trial aims to rigorously evaluate the efficacy of immersive VR aerobic exercise in enhancing exercise tolerance and patient satisfaction in hospitalized HF patients. Primary objective: To compare total exercise time between VR and standard cycle ergometer exercise. Secondary objectives: To assess between-group differences in perceived enjoyment (PACES)(18,19), and system usability (SUS)(20,21), as well as physiological responses.

### Explanation for the choice of comparator {9b}

The comparator is standard aerobic cycle-ergometer exercise, which represents the current clinical standard for inpatient cardiac rehabilitation at Hospital São Paulo and aligns with guideline-recommended practice for cardiac patients. By comparing immersive VR exercise to identical standard exercise (matching in duration, intensity prescription via modified Borg Scale, physiological monitoring, and supervision), this design isolates the specific effect of immersive VR technology while controlling for exercise intensity, therapeutic context, and clinician supervision.

### Rationale for active comparator design

This approach addresses a critical scientific question with direct clinical relevance: Does immersive VR technology enhance exercise tolerance and satisfaction beyond the established benefits of supervised aerobic exercise? By maintaining identical exercise parameters across groups, we isolate the VR-specific contribution while avoiding confounding from:

▯ Differential exercise intensities
▯ Unequal clinician attention
▯ Varying physiological monitoring procedures

This pragmatic, active comparator design maximizes clinical applicability and generalizability to practice settings where both immersive VR and standard exercise represent feasible rehabilitation options.

### Objectives {10}

#### Primary Objective

To evaluate the effect of immersive VR on exercise tolerance during cycle-ergometer aerobic exercise in hospitalized patients with HF.

#### Secondary Objectives

▯ To compare exercise enjoyment between groups using the Physical Activity Enjoyment Scale (PACES).
▯ To assess the usability of immersive VR with the System Usability Scale (SUS).
▯ To compare safety and physiological responses (adverse events, vital signs and VR-related symptoms) between groups.

## Methods: Patient and public involvement, and trial design

### Patient and public involvement {11}

This study was prospectively registered prior to the publication of the SPIRIT 2025 guidance, which introduced formal requirements for patient and public involvement reporting. Consequently, structured patient and public involvement was not incorporated into the original protocol design and registration.

However, the research question and study design were informed by extensive clinical experience with hospitalized heart failure patients and awareness of documented barriers to exercise-based rehabilitation participation.

Planned involvement in dissemination and future research phases:

- Following publication of primary results, we are committed to engaging patients and the public in disseminating and interpreting findings through the following activities:
- Lay summary and patient materials: A simplified summary of study methods, results, and implications will be developed in Portuguese and English for distribution to the public.
- Public seminars: Patient education seminars will be organized to present findings directly to heart failure patients, families, and caregivers.
- Healthcare provider communication: Findings will be disseminated to healthcare providers through professional conferences and publications.
- Future research collaboration: For longitudinal studies building on REVIVE-HF findings, formal patient and public involvement structures will be established during the design phase, including patient co-investigator roles where appropriate.

The research team recognizes the value of patient and public voices in health research and commits to transparent, accessible communication of results to all stakeholders.

### Trial design {12}

This study is a randomized, controlled, parallel-group clinical trial. Participants will be randomly allocated in a 1:1 ratio to either the control or immersive VR intervention group. Allocation will be conducted by an external researcher uninvolved in recruitment or intervention delivery, and data analysts will be blinded to group assignments. The trial investigates the effects of immersive VR on exercise tolerance in hospitalized heart failure patients, aiming to generate foundational evidence to support the use of immersive technologies in cardiac rehabilitation.

## Methods: Participants, interventions and outcomes

### Trial setting {13}

The study will be conducted on inpatient units in the Cardiology and Cardiac Surgery departments of Hospital São Paulo, São Paulo, Brazil. This referral hospital provides infrastructure for monitoring patients during interventions, including a qualified multidisciplinary team and all resources necessary to ensure participant safety and well-being.

### Superiority Design Justification

This trial uses a superiority design rather than a non-inferiority or equivalence design, based on the following rationale:

- Expected clinical advantage: The theoretical framework (attentional distraction, increased motivation) and preliminary data suggest immersive VR may provide benefits above standard exercise. The key research question is whether VR is superior to standard exercise for improving exercise tolerance.
- No defined non-inferiority margin: There is no established margin for what constitutes a clinically meaningful non-inferiority threshold for exercise tolerance in this clinical setting. Superiority trials are recommended when such margins are unavailable or unvalidated.
- Ethical and safety reasons: If immersive VR were inferior to standard exercise (i.e., associated with reduced exercise tolerance or safety issues), this finding would be critical for patient care and adoption decisions. Non-inferiority designs are more suitable once basic superiority and safety are clarified for novel interventions in vulnerable populations.

Should VR demonstrate superior outcomes, future research may consider non-inferiority or equivalence approaches for broader implementation or explore subgroup effects.

### Eligibility criteria for participants {14a}

#### Inclusion criteria

▯ Clinical diagnosis of HF with left ventricular ejection fraction (LVEF) ≤ 50% confirmed by echocardiography.
▯ Men and women aged ≥ 18 years.
▯ Hemodynamic stability for the previous 48 hours: no malignant ventricular arrhythmias; no complex extrasystoles; no supraventricular/sinus tachycardia > 120 bpm; no second- or third-degree atrioventricular block; and no signs of low output.
▯ New York Heart Association (NYHA) Functional Class I-IV (all functional classes included)
▯ Medical clearance for rehabilitation.
▯ Provision of written informed consent.

#### Non-inclusion criteria

▯ Arrhythmias that contraindicate physical exertion.
▯ Unstable angina.
▯ Acute myocardial infarction or cardiac surgery in the previous six months.
▯ Advanced or terminal cancer with life expectancy < 3 months (per physician)
▯ Cancer-related symptoms preventing safe exercise: severe fatigue, cachexia, uncontrolled pain (≥5/10 at rest), dyspnea at rest, or other cancer complications as determined by the investigator.

#### Exclusion Criteria

▯ Fever in the previous 48 hours, with leukocytosis and/or elevated C-reactive protein.
▯ Conditions limiting VR use, specifically: significant visual deficit, cognitive impairment, psychiatric or neurological disorders or history of photosensitive epilepsy.
▯ Orthopedic or neurological conditions precluding safe cycle-ergometer use.

### Eligibility criteria for sites and those delivering interventions {14b}

#### Study Site

This trial will be conducted at the Cardiology Division, Hospital São Paulo, Federal University of São Paulo (UNIFESP), São Paulo, Brazil.

#### Physiotherapist Qualification

The exercise intervention will be delivered by a licensed physiotherapist employed by Hospital São Paulo who is: registered with CREFITO (Regional Physical Therapy Council, Brazil), experienced in cardiac rehabilitation, and holds valid CPR certification.

#### Standardized Institutional Protocols

The exercise protocol and vital signs monitoring procedures are identical to institutional standard protocols already in routine use at Hospital São Paulo’s Cardiology Division. The physiotherapist will be trained and accustomed to: cycle ergometer operation, Modified Borg RPE assessment, vital signs measurement, electrocardiogram (ECG) monitoring, and standard exercise termination criteria. This standardization ensures consistency and minimizes variability in exercise delivery and monitoring across all study sessions.

#### VR-Specific Training

The only intervention difference between groups is the immersive VR addition. VR training includes: Meta Quest 2 operation, VZfit functionality, participant guidance, VR-specific safety procedures, and concurrent monitoring coordination. The physiotherapist will practice VR delivery before participant enrollment; training documentation will be maintained.

#### VR Protocol Standardization

The VR intervention will be delivered following standardized procedures to ensure consistency across all participants. The physiotherapist will receive training on VR delivery protocols, including proper equipment setup, participant guidance, and safety procedures. All VR sessions will adhere to a consistent protocol with standardized participant instructions and identical intervention parameters to minimize variability between sessions.

#### Data Collection Quality Control

Standardized data collection forms will be used for all measurements. Critical measurements will be recorded in real-time. Monthly team meetings will review exercise logs and data completeness; discrepancies will be discussed and addressed.

#### Study Supervision and Oversight

All interventions will be supervised by the Senior Investigator, Rita Simone Lopes Moreira, PhD.

### Who will take informed consent? {32a}

Informed consent will be obtained by trained members of the research team, specifically the Principal Investigator (Ariele dos Santos Costa, MSc) or designated research coordinators trained in Good Clinical Practice (GCP) and the study protocol.

The informed consent form will detail all relevant study information, including: trial objectives, procedures, expected duration of participant involvement, potential risks and benefits, confidentiality protections, and voluntary nature of participation. The form will be provided to participants (or their legal representatives if applicable) in Portuguese, allowing sufficient time for review and questions before written consent is obtained.

Participants will have the opportunity to ask questions and discuss any concerns with the research team before deciding whether to participate. Participation is entirely voluntary, and participants may withdraw at any time without affecting their standard clinical care or treatment at the institution.

All informed consent procedures will be conducted and overseen by the Research Ethics Committee of UNIFESP to ensure compliance with Brazilian regulations (CONEP- National Research Ethics Commission), the Declaration of Helsinki, and International Council for Harmonisation (ICH) guidelines for research involving human participants.

### Additional consent provisions for collection and use of participant data and biological specimens {32b}

No additional informed consent for ancillary studies is required. This trial does not include collection of biological specimens beyond clinical assessments performed as part of standard monitoring during the exercise session. Data collection is restricted to variables related to the intervention and monitoring of clinical and physical performance parameters (vital signs, perceived exertion, exercise duration). Participants will be informed during the initial consent process that data may be used for future research related to VR and cardiac rehabilitation; specific consent for future ancillary studies will be obtained separately if undertaken.

### Intervention and comparator description {15a}

#### Intervention Group: Immersive VR Cycle-Ergometer Exercise

##### Overview

Participants will complete a single supervised aerobic exercise session on a stationary cycle ergometer while wearing an immersive VR headset (Meta Quest 2), providing real-time visual and audio feedback synchronized with pedaling.

##### Exercise Protocol

- Equipment: Standard hospital cycle ergometer (no external load applied)
- Duration: Up to 20 minutes total (5 × 3-minute active pedaling blocks + 1-minute rest intervals) follows the published protocol described by Forestier et al (22), and has since become the standard of care at our institution for inpatient cardiac rehabilitation.
- Intensity: Self-selected cadence without RPE-guided prescription (target RPE 3–4 on Modified Borg Scale 0–10)
- Positioning: Seated upright in hospital chair
- Monitoring: Vital signs every minute (systolic/diastolic BP, HR, RR, SpO□), RPE every minute, real-time pedal cadence via VZfit

##### VR Environment

- Application: VZfit (VirZOOM Inc.) for Meta Quest 2
- Standardized Route: Pre-recorded scenic outdoor cycling route (countryside landscape with gentle terrain variation). All intervention participants experience the identical VR route to minimize environmental preference variability
- Route Options: Same scenic route available in daytime or nighttime ambiance
- Synchronization: Virtual avatar movement synchronized in real-time with participant pedaling speed; participants observe avatar progress through the virtual landscape
- Audio: Ambient nature sounds (wind, birds chirping, nature soundscape) corresponding to virtual setting; no music to avoid independent motivational confounding
- Interactivity: Virtual environment responds to pedaling cadence, providing real-time visual feedback of progress

##### VR Familiarization

1. Headset fitting and display adjustment (∼1 min): Proper fit, comfort verification, interpupillary distance (IPD) calibration, head strap tension
2. Orientation to virtual environment (∼2 min): Participant views VR environment while stationary; guided tour of the virtual setting
3. Movement practice (∼30–60 sec): Participant pedals at low intensity while observing synchronized avatar movement
4. Comfort and safety assessment: Supervising physiotherapist assesses for any discomfort, dizziness, or nausea; session proceeds only if participant comfortable
5. Safety procedures reviewed: Emergency exit procedures, communication methods, and participant controls explained

Participants experiencing significant discomfort during familiarization will not proceed to the exercise protocol.

##### Session Delivery

- Delivered by: Trained physiotherapist employed at Hospital São Paulo
- Timing: Daytime hours (08:00–16:00); consistent time of day for each participant to minimize circadian variability
- Single session: Only one exercise session per participant
- Real-time monitoring: Continuous observation for symptoms or hemodynamic changes
- Measurement timing: Vital signs and RPE recorded every minute during exercise and at recovery

#### Control Group: Standard Cycle-Ergometer Exercise

##### Overview

Participants will complete an identical exercise protocol to the intervention group, with the exception that no VR headset will be worn. Exercise occurs in the standard hospital room environment.

##### Exercise Protocol

###### Identical to intervention group

- Same cycle ergometer equipment and resistance settings
- Same exercise structure (5 × 3-minute active blocks, 1-minute rest intervals)
- Same total duration (up to 20 minutes)
- Same intensity prescription (self-selected cadence, RPE-guided, target RPE 3–4)
- Same monitoring procedures (vital signs every minute, RPE every minute, real-time cadence recording)
- Same session termination criteria
- Same physiotherapist supervision

###### Differences from intervention group

⊠ NO VR headset worn
⊠ Standard hospital room environment (windows, walls, standard ambient hospital sounds)
⊠ Standard cycle ergometer display showing time, cadence, estimated work (no virtual avatar or scenic environment visual)

##### Session Delivery

⊠ Delivered by: Trained physiotherapist
⊠ Timing: Same time of day and conditions as intervention group
⊠ Real-time monitoring: Identical vital signs, RPE, and observation protocols
⊠ Termination criteria: Identical to intervention group
⊠ Post-exercise: Identical to intervention group (recovery, assessment, discharge)

#### Only difference: Presence or absence of immersive VR technology

This active comparator design isolates the specific effect of VR immersion while controlling for all other exercise-related variables, allowing any observed differences in primary and secondary outcomes to be validly attributed to the VR intervention itself rather than differences in exercise prescription, supervision, or monitoring.

#### Single-Session Design Rationale

Single-session design was chosen for scientific and pragmatic reasons: (1) permits precise control over confounding variables (circadian effects, medication timing, rapidly changing clinical status); (2) demonstrates immediate physiological feasibility in a controlled setting, essential groundwork for future multi-center longitudinal studies; (3) minimizes resource demands in acute care settings with limited VR availability.

**Figure 1.**
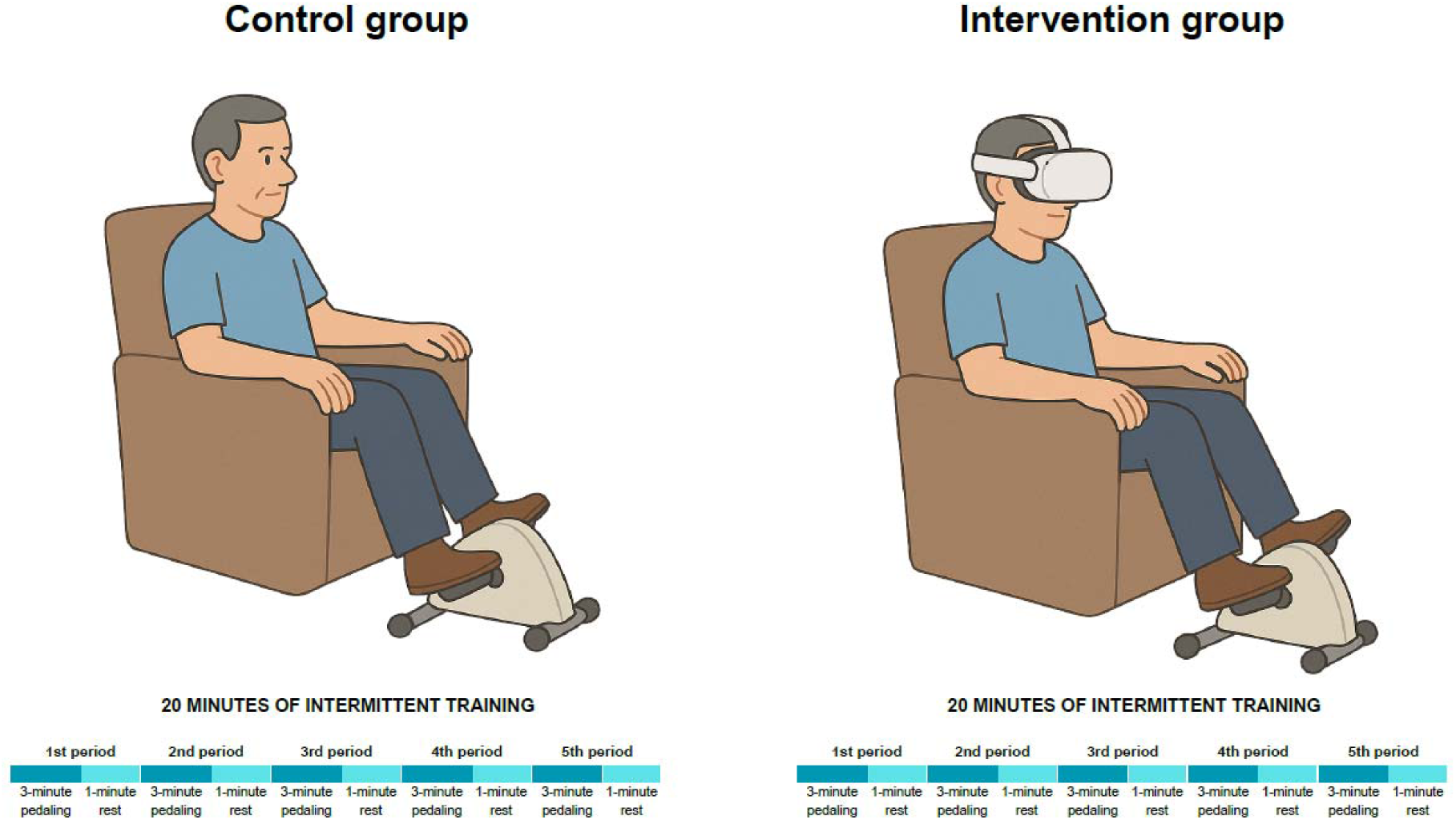
Schematic of the aerobic exercise protocol performed in the control and intervention groups.

### Criteria for discontinuing or modifying allocated intervention/comparator {15b}

Exercise will be discontinued immediately if any of the following occur:

#### Participant-Initiated Discontinuation

- Perceived dyspnea or lower-limb fatigue ≥ 5 on Modified Borg Scale (indicating excessive effort)
- Participant requests to stop due to discomfort, inability to continue, or any other reason
- Adverse effects related to the VR device compromising well-being, requiring suspension

#### Hemodynamic and Electrocardiographic Discontinuation

Participants will be continuously monitored using sphygmomanometer, pulse oximetry, ECG, and heart rate monitor. Exercise will be terminated immediately if any of the following occur, in accordance with the Brazilian Society of Cardiology’s 2024 Ergometry Consensus (23):

##### Blood Pressure abnormalities

- Diastolic blood pressure (DBP) > 120 mmHg in normotensive patients or > 140 mmHg in hypertensive patients
- Persistent drop in systolic blood pressure (SBP) ≥ 10 mmHg below baseline
- Marked rise in SBP > 200 mmHg

##### Heart Rate abnormalities

- Heart rate > 130 bpm or < 40 bpm
- Abnormal decrease in heart rate relative to exercise effort

##### Electrocardiographic abnormalities

- ST-segment depression ≥ 1 mm or ST-segment elevation
- Malignant arrhythmias (bigeminy, trigeminy, sustained ventricular tachycardia, atrial fibrillation with rapid ventricular response)

##### Clinical Manifestations

- Chest discomfort or pain
- Ataxia, dizziness, pallor, or cyanosis
- Presyncope or syncope
- Dyspnea disproportionate to effort (Modified Borg ≥ 5)

##### VR-System Issues (Intervention Group)

- Intolerance to the VR system compromising participant well-being (severe nausea, visual discomfort, severe dizziness)

##### Technical Failure

- Failure of monitoring systems (ECG, vital signs measurement) during intervention

##### Post-Discontinuation Protocol Upon immediate discontinuation

- Remove VR headset (intervention group only)
- Continue ECG monitoring for minimum 5 minutes
- Assess vital signs and symptoms
- Provide rest and recovery period
- Document reason for discontinuation

The study will take place in a hospital environment with immediate medical support. All sessions will be supervised by trained professionals to ensure safety and well-being throughout the intervention. These discontinuation criteria apply identically to both intervention and control groups.

### Strategies to improve adherence to intervention/comparator {15c}

Adherence is promoted through comprehensive participant engagement: detailed informed consent integrated into routine clinical care with emphasis on study contribution to cardiac rehabilitation science and minimal burden (single session). During exercise, continuous physiotherapist supervision provides real-time guidance, encouragement, and safety oversight. The VR group benefits from a structured 5-minute familiarization process minimizing technology anxiety, an immersive scenic virtual environment with synchronized avatar movement providing biofeedback.

Post-exercise, participants receive immediate feedback on total exercise time, positive reinforcement, and PACES/SUS questionnaires administered while experience is fresh. Study purpose is communicated emphasizing potential benefit to future heart failure patients.For this single-session design, adherence encompasses participant attendance and protocol completion while maintaining safety, comfort, and sense of agency.

### Concomitant care permitted or prohibited during the trial {15d}

Participants will be encouraged to maintain routine inpatient activities such as ambulation and sitting in an armchair, and to perform, twice daily, breathing exercises (deep inspiration and diaphragmatic breathing), active free limb exercises, and global stretching - standard unit interventions.

### Ancillary and post-trial care {34}

#### Immediately after study completion

Participants will continue to receive standard clinical care at Hospital São Paulo. Study participation consists of a single supervised exercise session and does not alter routine medical management. Following the exercise session, all participants will be encouraged to continue with standard inpatient cardiac rehabilitation.

#### After discharge from hospital

No specific study-related interventions beyond usual clinical care are planned after the study ends. Participants will follow routine post-hospitalization cardiac care in the outpatient setting.

#### Access to results

All participants will be informed about study results upon publication. Participants may request a summary of overall findings by contacting the research team.

#### Special provision for control group

Participants randomized to the control group (exercise without VR) will be offered an opportunity to experience the VR device after study completion, as a gesture of appreciation and to provide them access to the innovative intervention they did not receive during the study.

### Compensation for participation

Participants will not receive monetary compensation for study participation. All study procedures are provided at no cost to participants as part of routine inpatient care at Hospital São Paulo.

### Harm compensation

#### If adverse effects occur

If any participant experiences harm or adverse effects related to study participation, appropriate medical support will be provided immediately by Hospital São Paulo clinical staff. Any injury occurring during the study exercise session will be managed according to standard institutional protocols.

#### Injury compensation and liability

- Federal University of São Paulo (UNIFESP), as the sponsoring institution, maintains institutional liability coverage.
- Participants retain all legal rights under Brazilian law to seek compensation if harm occurs.
- Serious adverse events will be reported to the Research Ethics Committee for evaluation.

### Outcomes {16}

#### Primary outcome

- Influence of VR on tolerance to cycle-ergometer aerobic exercise in hospitalized patients with HF. The primary endpoint is effective exercise time (minutes), defined as total session duration until interruption due to physical limitations or symptoms, comparing VR vs. no-VR.

#### Secondary outcomes

1. Exercise time and clinical parameters (systolic/diastolic blood pressure, respiratory rate, peripheral oxygen saturation, heart rate, and rating of perceived exertion) during cycle-ergometer exercise with and without VR. These parameters will be measured across the exercise session, comparing the two approaches.
2. Usability of the VR system during exercise, assessed using the System Usability Scale (SUS) [11,12] administered after the session (intervention group): ease of use, comfort, and perceived immersion.
3. Patient satisfaction with the exercise experience, comparing sessions with and without VR, measured using the Physical Activity Enjoyment Scale (PACES) [13,14].

### Harms {17}

#### Anticipated harms and safety considerations

##### Exercise-related harms (applicable to both groups)

The primary anticipated harms relate to the physiological stress of exercise in hospitalized heart failure patients. These include:

###### 1. Cardiovascular events

▯ Acute coronary syndrome symptoms (chest pain, dyspnea, diaphoresis)
▯ Dangerous arrhythmias (sustained ventricular tachycardia, rapid atrial fibrillation)
▯ Hemodynamic instability (acute hypotension, cardiogenic shock)
▯ Syncope or presyncope

###### 2. Respiratory events

▯ Severe dyspnea
▯ Oxygen desaturation (SpO□ <85%)

###### 3. Musculoskeletal

▯ Muscle cramps
▯ Leg pain or exacerbation of joint pain

##### VR-specific harms (intervention group only)

###### 1. Cybersickness

▯ Nausea or vomiting
▯ Dizziness or vertigo
▯ Disorientation
▯ Headache or eye strain

###### 2. Device-related

▯ Skin irritation from headset contact
▯ Equipment malfunction preventing safe use

#### Definition and assessment of harms

Expected harms are defined as adverse events anticipated based on:

▯ Physiological response to exercise in HF patients
▯ Known characteristics of VR technology in similar populations
▯ Literature reporting on exercise stress testing and cardiac rehabilitation
▯ Unexpected harms are defined as adverse events not anticipated or not mentioned in this protocol.

All harms will be assessed systematically through:

1. Continuous monitoring during exercise:

▯ Real-time vital signs assessment (every minute)
▯ Continuous visual observation
▯ Verbal communication with participant
2. Standardized questionnaires:

▯ Modified Borg Rating of Perceived Exertion (0-10 scale) - every minute
▯ Post-exercise symptom assessment
▯ System Usability Scale (for VR-related issues in intervention group)
3. Adverse event documentation:

▯ Type of adverse event
▯ Timing (during exercise and during recovery)
▯ Severity classification (mild, moderate, severe)
▯ Relatedness to study procedures (definite, probable, possible, unrelated)
▯ Whether event required intervention/treatment

#### Harms classification and standardization

Severity classification:

▯ Mild: Uncomfortable but does not interfere with daily function; resolves spontaneously or with minimal intervention
▯ Moderate: Interferes with daily function or requires medical intervention; may persist
▯ Severe: Incapacitating; requires hospitalization or prevents exercise continuation; potentially life-threatening

Relatedness assessment:

▯ Definite: Clear temporal relationship with study procedure; no alternative explanation
▯ Probable: Temporal relationship consistent with study procedure; alternative explanation less likely
▯ Possible: Temporal relationship uncertain; alternative explanations possible
▯ Unrelated: Clear alternative explanation; temporal relationship inconsistent

Standardized language:

Harms will be documented using descriptive terminology aligned with common cardiovascular and respiratory symptom classifications. VR-specific symptoms will be documented using standard cybersickness terminology (nausea, dizziness, disorientation, headache, eye strain).

#### Reporting approach for adverse events

All serious adverse events will be reported, defined as events requiring hospitalization, causing persistent incapacity, resulting in permanent disability, or potentially life-threatening. All VR-related adverse events will be reported, defined as cybersickness symptoms (nausea, dizziness, disorientation, headache, eye strain) occurring during or immediately after study procedures. All exercise-stopping events will be reported, defined as any adverse event leading to early session termination before planned protocol completion. Adverse events occurring in ≥5% of participants in either group will be reported. Any adverse event causing permanent harm will be reported. Non-serious adverse events not meeting above criteria may be summarized descriptively but will not be listed individually in results.

#### Harms monitoring and safety procedures

##### Real-time monitoring during exercise

The supervising physiotherapist has authority to terminate the exercise session immediately if any safety concern arises. Medical support is available immediately if needed, with emergency equipment accessible per hospital protocol. Participants have the explicit right to request session termination at any time for any reason, without penalty or compromise to clinical care.

##### Post-session assessment

Vital signs (BP, HR, RR, SpO□) will be assessed at 1 minute post-exercise and monitored continuously until participant hemodynamic stability is confirmed (typically 5 minutes). Participants will be instructed to report any delayed adverse symptoms (occurring >1 hour after exercise completion) within 24 hours via telephone contact with research team. Contact information will be provided to all participants before exercise begins.

##### Serious adverse event reporting

Any serious adverse event (as defined in Reporting Approach section) occurring during or within 24 hours post-exercise will be reported to the Principal Investigator immediately (within 2 hours of identification). Serious adverse events will be reported to the Research Ethics Committee of UNIFESP within 48 hours. All events will be assessed for temporal and causal relationship to study procedures using standardized relatedness categories (definite, probable, possible, unrelated). If a pattern of concerning adverse events emerges (≥2 serious events potentially related to study procedures), the Principal Investigator will assess whether Data Safety Monitoring Board review or study pause is warranted.

#### Data analysis approach for harms

Adverse event rates (number of events, number of participants experiencing events) will be compared between the immersive VR intervention group and the standard exercise control group using chi-square tests for between-group differences in event incidence. Between-group differences in adverse event rates will be examined, with particular attention to VR-specific harms (cybersickness symptoms). Within-group adverse event trends will be monitored during recruitment, with events documented by participant sequence and group assignment to identify any temporal patterns suggesting emerging safety signals. Any suspected safety signals (unexpected increase in serious adverse events, or higher-than-anticipated adverse event rate in one group) will be reviewed by the research team and discussed with the Ethics Committee for determination of appropriate action (e.g., protocol modification, participant notification, study pause, or continuation with enhanced monitoring).

### Participant timeline {18}

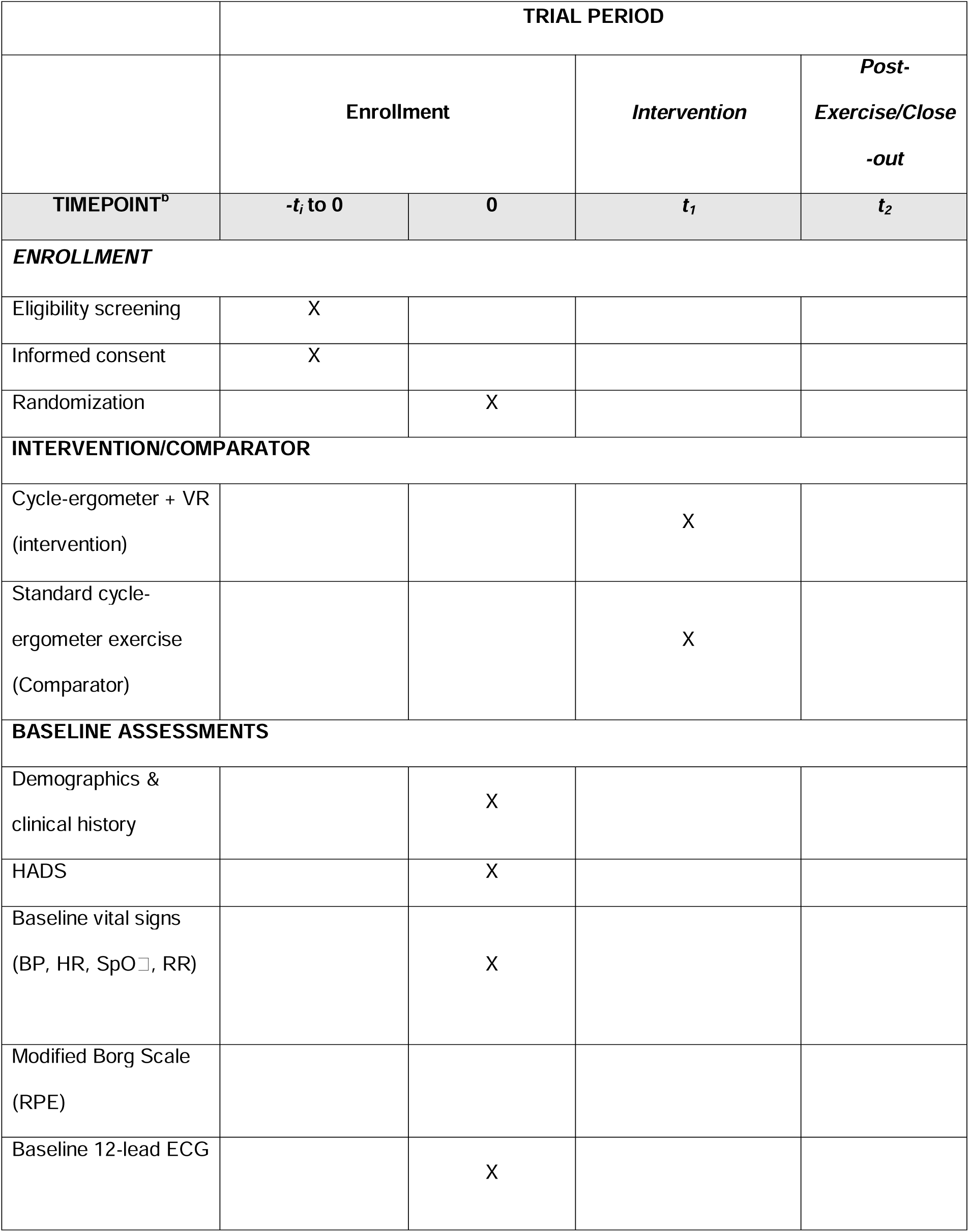
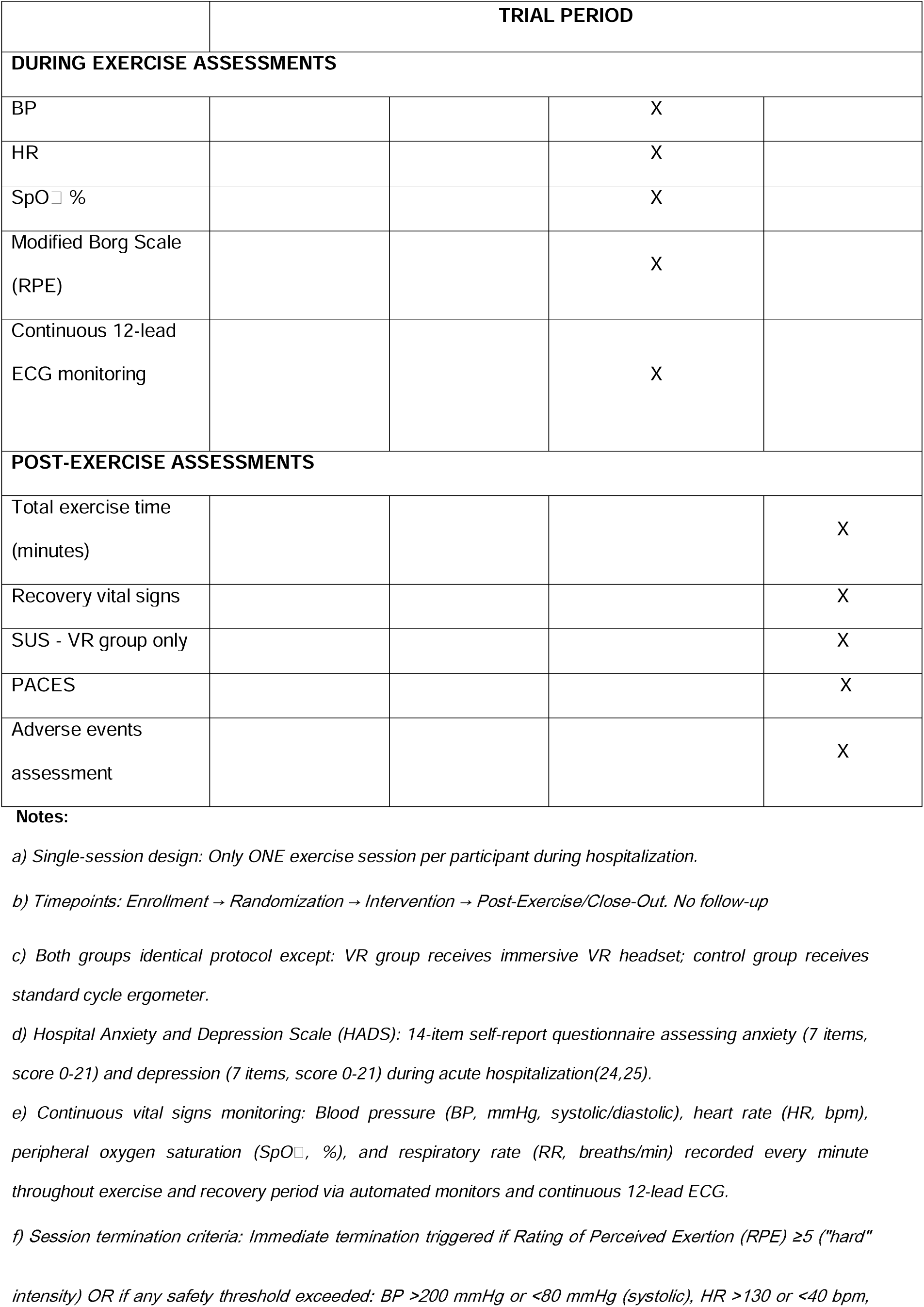
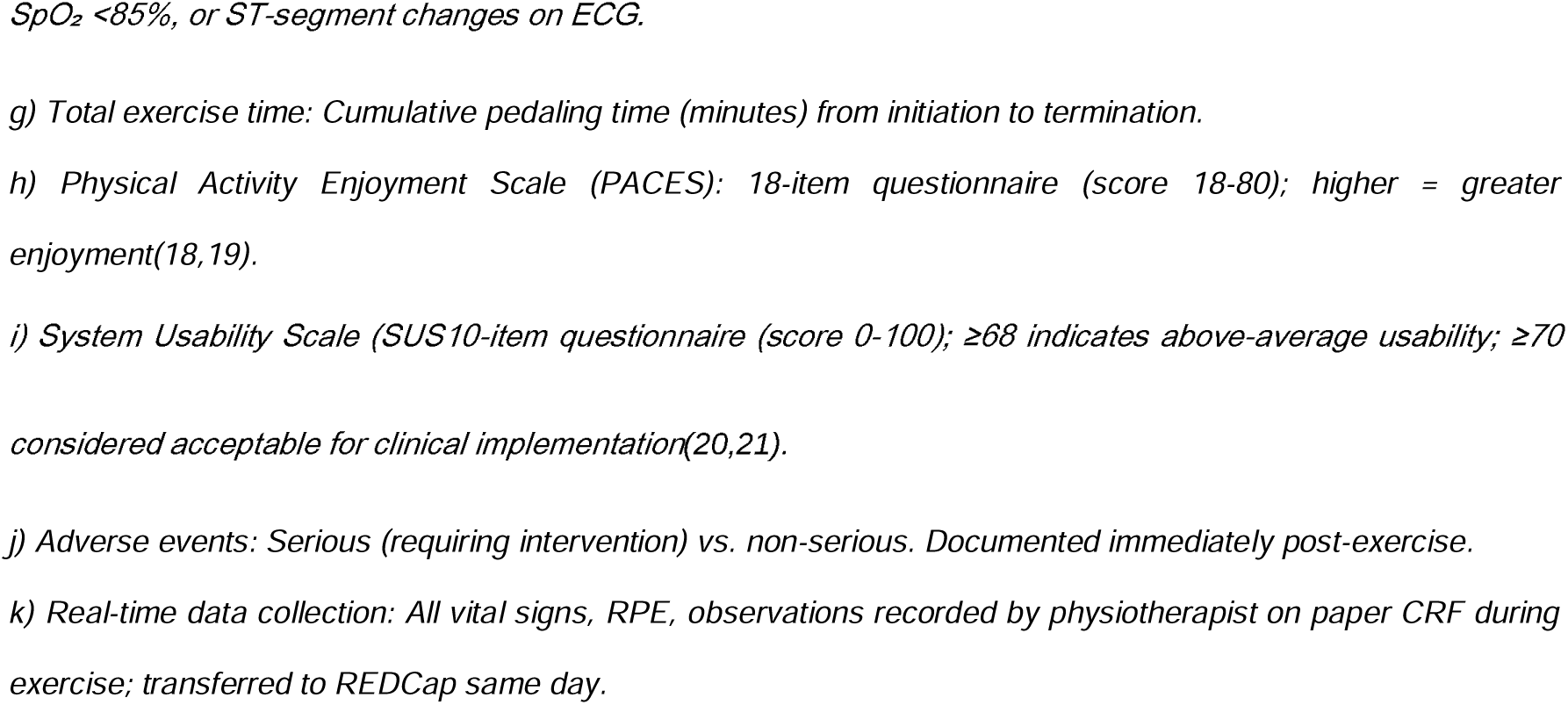

|  | TRIAL PERIOD |  |  |  |
| --- | --- | --- | --- | --- |
|  | Enrollment |  | Intervention | Post-Exercise/Close<br>-out |
| TIMEPOINT<sup>b</sup> | -$t_i$ to 0 | 0 | $t_1$ | $t_2$ |
| **ENROLLMENT** |  |  |  |  |
| Eligibility screening | X |  |  |  |
| Informed consent | X |  |  |  |
| Randomization |  | X |  |  |
| **INTERVENTION/COMPARATOR** |  |  |  |  |
| Cycle-ergometer + VR<br>(intervention) |  |  | X |  |
| Standard cycle-ergometer exercise<br>(Comparator) |  |  | X |  |
| **BASELINE ASSESSMENTS** |  |  |  |  |
| Demographics &<br>clinical history |  | X |  |  |
| HADS |  | X |  |  |
| Baseline vital signs<br>(BP, HR, SpO<sub>2</sub>, RR) |  | X |  |  |
| Modified Borg Scale<br>(RPE) |  |  |  |  |
| Baseline 12-lead ECG |  | X |  |  |
|  | TRIAL PERIOD |  |  |  |
| DURING EXERCISE ASSESSMENTS |  |  |  |  |
| BP |  |  | X |  |
| HR |  |  | X |  |
| SpO<sub>2</sub> % |  |  | X |  |
| Modified Borg Scale<br>(RPE) |  |  | X |  |
| Continuous 12-lead<br>ECG monitoring |  |  | X |  |
| POST-EXERCISE ASSESSMENTS |  |  |  |  |
| Total exercise time<br>(minutes) |  |  |  | X |
| Recovery vital signs |  |  |  | X |
| SUS - VR group only |  |  |  | X |
| PACES |  |  |  | X |
| Adverse events<br>assessment |  |  |  | X |

**Notes:**
- a) Single-session design: Only ONE exercise session per participant during hospitalization. - b) Timepoints: Enrollment → Randomization → Intervention → Post-Exercise/Close-Out. No follow-up - c) Both groups identical protocol except: VR group receives immersive VR headset; control group receives standard cycle ergometer. - d) Hospital Anxiety and Depression Scale (HADS): 14-item self-report questionnaire assessing anxiety (7 items, score 0-21) and depression (7 items, score 0-21) during acute hospitalization(24,25). - e) Continuous vital signs monitoring: Blood pressure (BP, mmHg, systolic/diastolic), heart rate (HR, bpm), peripheral oxygen saturation (SpO<sub>2</sub>, %), and respiratory rate (RR, breaths/min) recorded every minute throughout exercise and recovery period via automated monitors and continuous 12-lead ECG. - f) Session termination criteria: Immediate termination triggered if Rating of Perceived Exertion (RPE) ≥5 ("hard" intensity) OR if any safety threshold exceeded: BP >200 mmHg or <80 mmHg (systolic), HR >130 or <40 bpm,
*SpO<sub>2</sub> <85%, or ST-segment changes on ECG.*
*g) Total exercise time: Cumulative pedaling time (minutes) from initiation to termination.*
*h) Physical Activity Enjoyment Scale (PACES): 18-item questionnaire (score 18-80); higher = greater enjoyment(18,19).*
*i) System Usability Scale (SUS 10-item questionnaire (score 0-100); ≥68 indicates above-average usability; ≥70 considered acceptable for clinical implementation(20,21).*
*j) Adverse events: Serious (requiring intervention) vs. non-serious. Documented immediately post-exercise.*
*k) Real-time data collection: All vital signs, RPE, observations recorded by physiotherapist on paper CRF during exercise; transferred to REDCap same day.*

### Sample size {19}

The sample size was estimated for adult inpatients with clinical HF decompensation who are hemodynamically stable and meet the study’s eligibility criteria. This is a prospective, randomized trial with a total sample of 60 participants (30 per group, control and intervention), defined by computer-generated 1:1 with no stratified.

Calculations were performed in G*Power (v3.1.9.2) assuming a superiority design with an independent-samples t-test as the primary analysis. Based on prior VR exercise studies (11,17,26), we assumed a moderate effect size (Cohen’s d = 0.65) for the primary outcome (total exercise duration completed). With alpha set at 0.05 (two-tailed), beta at 0.20 (80% power), and assuming 10% dropout, the required sample size was determined to be 30 participants per group (n = 60 total). Secondary per-protocol analysis will include participants who completed the entire exercise session without unplanned termination.

### Recruitment {20}

Participants will be identified through medical record review based on inclusion criteria. After preselection, patients will be invited to participate, receive detailed information about objectives and procedures, and provide written informed consent. A complete clinical assessment will follow, including demographic/sociodemographic data, history, medications, key complementary exams, and physical examination. The Hospital Anxiety and Depression Scale (HADS) (24,25) will be administered. Participants will then be randomized to intervention or control.

### Assignment of interventions: randomization

#### Sequence generation: who will generate the sequence {21a}

The allocation sequence will be generated using a computerized random number generator within the secure REDCap (Research Electronic Data Capture) platform. The sequence will be prepared by an external biostatistician who is not involved in patient recruitment or study procedures. **Sequence generation: type of randomization {21b}**

Participants will be randomized individually using a computer-generated block randomization sequence with a 1:1 allocation ratio to intervention or control group. Block randomization (fixed block size of 4) ensures balanced group sizes throughout the trial. No stratification will be used. Only the statistician responsible for sequence generation will have access prior to study start.

#### Allocation concealment mechanism {22}

Participant enrollment will be performed by the research team, who will conduct screening, data collection, and study explanations. After consent, the clinical assessment will be performed, including HADS administration.

#### Implementation {23}

Assignment to interventions will be carried out by an external biostatistician using a pre-generated randomization sequence, ensuring impartial and concealed allocation to control or VR intervention groups. Randomization will occur after informed consent is obtained and baseline assessments are completed, preventing selection bias. The biostatistician responsible for randomization sequence generation, concealment, and allocation has no direct contact with study participants, no involvement in participant recruitment or eligibility screening, no role in intervention delivery or supervision, and no access to outcome data during trial conduct. This individual’s sole responsibility is generating the randomization sequence using appropriate methodology (block randomization with 1:1 allocation ratio), maintaining allocation concealment via secure electronic system, and revealing allocation assignments only after participant informed consent and baseline assessment completion.

All randomization activities are conducted remotely via secure electronic communication (REDCap platform with restricted access). Participants and trial staff will not be able to predict or influence allocation assignments, as the randomization sequence is generated remotely and revealed only at the designated time. Physical separation of randomization personnel from clinical trial conduct ensures complete independence and minimizes risk of selection bias or allocation manipulation. The external biostatistician will maintain the randomization sequence in a secure.

### Assignment of interventions: blinding

#### Who will be blinded {24a}

##### Blinding Limitations

Due to the nature of the VR intervention, participants and the physiotherapist cannot be blinded. Participants will necessarily be aware of whether they are receiving standard exercise or VR-augmented exercise, as the presence or absence of the immersive VR headset is inherently visible. Similarly, the physiotherapist delivering the intervention must know group allocation to deliver the assigned intervention appropriately.

##### Measures to Minimize Bias

Despite the lack of participant and intervention provider blinding, bias will be minimized through the following strategies:

▯ **Objective Measurement Standardization:** Vital signs and exercise duration will be recorded using calibrated, automated monitoring equipment in real-time, reducing observer bias in physiological measurements.
▯ **Standardized Procedures:** All participants in both groups will receive identical exercise protocols, duration, intensity monitoring procedures, and standardized verbal instructions, ensuring that measurement procedures are equivalent across groups.
▯ **Data Analyst Blinding:** Statistical analyses will be conducted by an external biostatistician using blinded data coding (Group A/Group B only), without knowledge of intervention group assignment during analysis to prevent analysis bias.
▯ **Allocation Concealment:** Participants and clinical staff will not know group assignment until after baseline assessments are completed.

#### How will be blinding be achieved {24b}

▯ **Data Analyst:** Blinded to group allocation during statistical analysis
▯ **External Randomization Manager:** Maintains allocation concealment until baseline assessments are complete
▯ **Participants and Physiotherapist:** Cannot be blinded due to intervention visibility, but will not be informed of trial hypotheses or expected outcomes

The data analyst will remain blinded to group allocation until completion of all primary statistical analyses and generation of the primary results table. Group identities will be revealed to the analyst only after the primary analyses have been completed, documented, and verified.

#### Procedure for unblinding if needed {24c}

If it becomes necessary to reveal a participant’s allocation, unblinding will occur in a controlled manner. The person responsible for randomization and case oversight may access the participant’s assignment through a secure system without sharing that information with other team members or influencing outcome assessments. All unblinding decisions will be documented with justification and context.

### Data collection and management

#### Plans for assessment and collection of outcomes {25a}

Data will be collected by trained physiotherapist who will supervise all exercise sessions and record data using standardized case report forms (CRF). Patient-reported satisfaction with the exercise experience will be assessed using the Physical Activity Enjoyment Scale (PACES)(18,19), a 18-item self-report questionnaire administered immediately post-exercise to both groups. For the VR intervention group, system usability will be assessed using the System Usability Scale (SUS)(20,21), a 10-item questionnaire administered immediately post-exercise to measure ease of use, comfort, and perceived immersion with the VR technology.

Vital signs monitoring will be performed before, during (every minute), and after exercise using standardized equipment: pulse oximetry for oxygen saturation (SpO□), automated sphygmomanometer for systolic and diastolic blood pressure (BP), and continuous heart rate monitoring via pulse oximeter and ECG. Continuous 12-lead ECG monitoring via bedside cardiac monitor will be performed during all exercise sessions for safety surveillance. Continuous ECG waveforms during the exercise session will not be archived. Only ECG strips corresponding to detected clinically significant arrhythmias (atrial fibrillation, ≥6 consecutive ventricular ectopic beats, sustained ventricular tachycardia >3 beats) or ischemic changes (≥1 mm ST-segment deviation from baseline) will be saved for post-hoc review by a blinded cardiologist.

Perceived exertion will be assessed using the Modified Borg Rating of Perceived Exertion Scale (0–10), administered at baseline, every minute during exercise, and post-exercise to monitor exercise intensity and participant tolerance. All outcome data will be recorded in real-time on standardized forms during the exercise session to minimize measurement error and recall bias. Post-exercise questionnaires (PACES and SUS) will be administered while participant experience remains fresh, typically 5–10 minutes after exercise completion. All data will be entered into the secure REDCap database with mandatory quality checks for completeness, consistency, and integrity before database lock.

#### Plans to promote participant retention and complete follow-up {25b}

1. **Continuous monitoring:** During hospitalization, patients will be supported by a multidisciplinary team (physician, nutritionist, pharmacist, psychologist, physiotherapist, and nurse).
2. **Active engagement:** Before starting, participants will be informed about the benefits of the intervention; VR will be presented as an innovative tool to enhance the experience.
3. **Personalized support:** The team will offer ongoing assistance and address questions; participants may interrupt or modify the intervention if needed without jeopardizing care.
4. **Detailed records:** All deviations/interruptions will be recorded; participants who withdraw will be followed to document reasons and mitigate impact.
5. **Clear communication:** From admission to discharge, the study’s importance and participants’ contribution will be emphasized.

#### Data management {26}

▯ Data entry: All data will be entered electronically into REDCap using standardized forms covering sociodemographic, clinical data, monitoring parameters, perceived exertion, and questionnaires.
▯ Coding: Participants will be identified by randomly generated codes to ensure anonymity; no personally identifiable information will be linked to analytic datasets.
▯ Security: Access will be restricted to authorized researchers with role-based permissions; data will be encrypted within REDCap.
▯ Archiving: After completion, data will be archived for at least 5 years per ethical/regulatory guidance, with restricted access.
▯ Quality assurance: Range checks and consistency reviews will be applied; all data collectors will receive training to ensure standardized, reliable measurements.

#### Confidentiality {33}

**Collection of personal information:** Personal and clinical data (e.g., sociodemographic, medical history, medications, exam results) will be collected solely for research purposes with participant authorization via written informed consent.

##### Protection of confidentiality

▯ De-identification: Participants will be identified by unique codes; personally identifiable information will be stored separately from study data.
▯ Secure storage: Data will be stored on secure platforms (e.g., REDCap) with end-to-end encryption; access will be limited to authorized team members and to the minimum necessary.
▯ Restricted access: Password-protected accounts will limit data viewing to those involved in collection/analysis.
▯ Sharing: Information will be used exclusively for study purposes and will not be shared with third parties without prior consent. Any required sharing (e.g., for auditing) will occur only in aggregate and de-identified form.
▯ Post-study: Data will be securely maintained for a minimum of 5 years; access will remain restricted; secure disposal will follow expiration of the retention period.
▯ Participant assurances: All procedures will be explained in the informed consent, and participants may request deletion of their personal information as allowed by ethical regulations.

### Statistical methods

#### Statistical methods for primary and secondary outcomes {27a}

Analyses will be performed using SPSS Statistics v25.0 with significance level α = 0.05 (two-tailed). Primary outcome (total exercise duration) will be compared between groups using independent-samples t-test, or Mann-Whitney U test if normality is violated (Shapiro-Wilk p < 0.05). Secondary outcomes will be compared using appropriate parametric or non-parametric tests. Likert-scale items will be analyzed using Mann-Whitney U tests. Descriptive statistics (means ± SD, medians [IQR]) will characterize all outcomes.

#### Who will be included in each analysis {27b}

All randomized participants who provided informed consent and were allocated to a group will be included in the primary intention-to-treat (ITT) analysis, according to their original allocation, regardless of adherence or protocol deviations. Secondary per-protocol analysis will include participants who completed the entire planned exercise session without unplanned termination due to participant request or safety concern. Analysis will be based on actual exercise duration achieved by each participant (measured continuously, independent of exercise structure). Results from per-protocol analysis will be compared with intention-to-treat findings to assess robustness of primary findings.

#### How missing data will be handled in the analysis {27c}

For primary and secondary outcomes, every effort will be made to minimize missing data through robust follow-up and documentation procedures during the single-session design. If missing data occur for the primary outcome, a complete-case analysis will be performed. As a sensitivity analysis, multiple imputation will be considered if the proportion of missing data exceeds 5%. Missing data patterns and reasons will be described in detail. All analyses involving missing data will be clearly reported, and no participants will be excluded solely due to missing data for secondary or exploratory outcomes.

#### Methods for additional analyses (e.g. subgroup analyses) {27d}

Pre-specified subgroup analyses will examine potential treatment effect modifications by age, sex, LVEF and NYHA functional class. Subgroup-by-treatment interactions will be tested using linear regression with group-by-variable interaction terms. The significance threshold for interactions is α = 0.10 (exploratory). Results should be interpreted cautiously given potential for Type I error. Multivariable linear regression models will be used to adjust outcomes and control for potential confounders in secondary analyses.

#### Interim analyses {28b}

No interim analyses are planned. Any decision to interrupt the study will be based on relevant clinical or ethical criteria (e.g., serious adverse events). If necessary, interim analyses will be performed exclusively by an independent data monitoring entity with restricted access to partial results.

#### Protocol and statistical analysis plan {5}

The full protocol, participant-level data, and statistical code will be made available upon reasonable request in accordance with institutional policies. Access will be controlled to protect privacy and confidentiality and will require ethics approval. Documentation and code will be hosted on the university’s institutional data repository with appropriate safeguards.

### Oversight and monitoring

#### Composition of the coordinating centre and trial steering committee {3d}

A formal Trial Steering Committee (TSC) or Outcomes Adjudication Committee was not deemed necessary; coordination will be centralized within the core research team (cardiology specialists). The principal investigator will oversee study conduct and ethical decisions. Data collection, storage, and analysis will be managed by an external researcher who will perform regular quality and compliance checks. All ethical/regulatory aspects will be supervised by the institutional Research Ethics Committee. Participant safety monitoring will follow good clinical practice and established guidelines; no additional data monitoring or adjudication committee is planned.

#### Composition of the data monitoring committee, its role and reporting structure {28a}

No Data Monitoring Committee (DMC) is planned given the low-risk intervention and continuous oversight by experienced professionals. Participant safety will be regularly monitored by the main research team. Any serious adverse event will prompt appropriate action in accordance with institutional safety/ethics guidance. Safety oversight will be supervised by the institutional Research Ethics Committee.

#### Frequency and plans for auditing trial conduct {29}

Independent audits will be conducted periodically to ensure compliance with the protocol and good research practices. At minimum, two audits will occur over the course of the study to ensure data quality, integrity, and early identification of deviations or inconsistencies.

#### Protocol amendments {31}

Any substantial amendment - including changes in eligibility criteria, objectives, analyses, or interventions - will be submitted for approval to the Research Ethics Committee (REC) and communicated to investigators via meetings or formal notices. If modifications directly affect participants, they will be informed and asked to sign an updated consent form, if needed. Amendments will also be recorded in the clinical trials registry and reported in scientific outlets to ensure transparency and communicated to regulatory authorities as required.

Any substantial protocol modifications will follow this procedure:

1. Amendment drafted by Principal Investigator with scientific justification
2. Ethics Committee approval required before implementation (UNIFESP CEP)
3. ReBEC registry update within 2 weeks of approval
4. Investigator notification via formal meeting or written notice
5. Participant notification if changes affect inclusion/procedures
6. Trial registry update (ensaioclinico.gov.br) if applicable

#### Dissemination policy {8}

Results will be reported according to CONSORT guidelines and published in peer-reviewed journals and international conferences. Findings will be disseminated to patient advocacy organizations for heart failure, clinical guideline committees, and professional societies to inform evidence-based cardiac rehabilitation protocols. De-identified data will be deposited in publicly accessible research databases to enable transparency and secondary analyses.

## Discussion

This study addresses a relevant gap in inpatient cardiac rehabilitation by exploring the use of VR to enhance tolerance to aerobic exercise among hospitalized patients with HF. We hypothesize that VR during exercise may increase engagement, improve patient experience, and favor adherence to rehabilitation programs even in hospital settings.

Practical and operational challenges typical of hospital environments are anticipated, such as resource availability, integration with routine care, and logistics to schedule exercise sessions without disrupting clinical activities. Rigorous protocol adherence and precise data collection require continual communication between the research team and clinical staff. Although innovative, VR requires adequate training to ensure safe and effective use and to minimize technical issues or patient discomfort.

Critical aspects include tight safety monitoring given the fragile clinical condition of patients with HF and the protection of confidentiality in accordance with ethical and regulatory standards. These challenges will be mitigated through structured safety protocols, trained personnel, and continuous supervision during the intervention.

### Anticipated Challenges and Mitigation

Hospital-based implementation requires coordination with clinical workflows, staff training in VR technology, and tight safety monitoring. These challenges will be mitigated through: (1) structured safety protocols with predefined termination criteria; (2) comprehensive staff training; (3) real-time clinical supervision; and (4) secure REDCap data management per Brazilian regulatory standards (CONEP).

### Future Directions and Expected Outcomes

Our pilot study(17) demonstrated feasibility of immersive VR exercise in hospitalized HF patients. This REVIVE-HF trial represents a rigorous advancement with adequate sample size, randomized design, and predefined outcomes. We expect the VR group to achieve longer exercise duration and higher enjoyment (PACES) and usability (SUS) scores compared to controls. If confirmed, this protocol could be adapted to other hospital contexts and incorporated as a complementary resource in cardiac rehabilitation programs, contributing to innovation in acute care.

#### Trial status

**Protocol version:** 1.0; Date: 26 October 2025

▯ **Recruitment start date:** 05 January 2026
▯ **Anticipated end of recruitment:** 05 December 2026

## Data Availability

All data generated in this trial will be made available in de-identified form in an open-access repository (e.g., Zenodo) upon completion of the study and reasonable request to the corresponding author, in accordance with institutional and ethics committee policies.

## Abbreviations

AV block: Atrioventricular block
bpm: Beats Per Minute
CAAE: Certificado de Apresentação para Apreciação Ética
CEP: Research Ethics Committee
CONSORT: Consolidated Standards of Reporting Trials
CONEP: National Research Ethics Commission
CPR: Cardiopulmonary Resuscitation
CR: Cardiac Rehabilitation
CREFITO: Regional Physical Therapy Council, Brazil
CRF: Case Report Form
CSV: Comma-Separated Values
CVD: Cardiovascular Diseases
DBP: Diastolic Blood Pressure
DMC: Data Monitoring Committee
DOI: Digital Object Identifier
ECG: Electrocardiogram
GCP: Good Clinical Practice
GDPR: General Data Protection Regulation
G*Power: Statistical Power Analysis Software
HF: Heart Failure
HR: Heart Rate
HADS: Hospital Anxiety and Depression Scale
IBGE: Brazilian Institute of Geography and Statistics
IPD: Individual Participant Data;
ID: Interpupillary Distance
ITT: Intention-to-Treat
LVEF: Left Ventricular Ejection Fraction
mHealth: Mobile Health
min: Minutes
mmHg: Millimeters of Mercury
NYHA: New York Heart Association
PACES: Physical Activity Enjoyment Scale
RCT: Randomized Controlled Trial
ReBEC: Brazilian Clinical Trials Registry
REDCap: Research Electronic Data Capture
RPE: Rating of Perceived Exertion
RPM: Revolutions Per Minute
RR: Respiratory Rate
SBP: Systolic Blood Pressure
SOP: Standard Operating Procedure
SpO□: Peripheral Oxygen Saturation
SPIRIT: Standard Protocol
Items: Recommendations for Interventional Trials
SPSS: Statistical Package for the Social Sciences
SUS: System Usability Scale
SVT: Supraventricular Tachycardia
TSC: Trial Steering Committee
UNIFESP: Federal University of São Paulo
VR: Virtual Reality
VZfit: VirZOOM Immersive Exercise Application
W: Watts
WHO: World Health Organization

## Declarations Acknowledgements

The authors thank the GAEPIIS Research Group in Innovation and Health for technical and intellectual support throughout the study’s development.

## Authors’ contributions {3a}

▯ **ASC -** Study conception, methodology, and manuscript writing.
▯ **CBB** - Study conception, methodology, critical protocol review, and methodological support.
▯ **SG** - Scientific supervision, critical protocol review, and methodological support.
▯ **IBK** - Scientific supervision, critical protocol review, and protocol validation.
▯ **PIMM** - Scientific supervision, critical protocol review, and protocol validation.
▯ **VRS** - Study conception, methodology, critical protocol review, and methodological support.
▯ **RSLM** - Overall study coordination and final manuscript review. All authors read and approved the final manuscript.

## Availability of data and materials {6}

The final dataset will be available from the corresponding author upon reasonable request. There are no contractual agreements limiting investigator access.

## Ethics approval and consent to participate {30}

The study will be conducted in accordance with international ethical standards and was approved by the Research Ethics Committee of the Universidade Federal de São Paulo (UNIFESP) under approval number [CAAE: 54896621.9.0000.5505]. Written informed consent will be obtained from all participants before enrollment. Participants will be informed about study objectives, procedures, potential risks and benefits, and their right to withdraw at any time without prejudice.

## Consent for publication

Not applicable, as no identifiable personal data, images, or videos will be included. If necessary, a consent model for publication of participant-related details will be available upon request.

## Competing interests {7b}

The authors declare no competing interests.

## References

1. Groenewegen A, Rutten FH, Mosterd A, Hoes AW. Epidemiology of heart failure. Eur J Heart Fail. 2020 Aug;22(8):1342–56.

2. Bocchi EA, Arias A, Verdejo H, Diez M, Gómez E, Castro P. The Reality of Heart Failure in Latin America. J Am Coll Cardiol. 2013 Sep;62(11):949–58.

3. Heidenreich PA, Bozkurt B, Aguilar D, Allen LA, Byun JJ, Colvin MM, et al. 2022 AHA/ACC/HFSA Guideline for the Management of Heart Failure: A Report of the American College of Cardiology/American Heart Association Joint Committee on Clinical Practice Guidelines. Circulation. 2022 May 3;145(18).

4. Taylor RS, Walker S, Ciani O, Warren F, Smart NA, Piepoli M, et al. Exercise-based cardiac rehabilitation for chronic heart failure: the EXTRAMATCH II individual participant data meta-analysis. Health Technol Assess (Rockv). 2019 May;23(25):1–98.

5. Taylor RS, Dalal HM, Zwisler AD. Cardiac rehabilitation for heart failure: ‘Cinderella’ or evidence-based pillar of care? Eur Heart J. 2023 May 1;44(17):1511–8.

6. Chindhy S, Taub PR, Lavie CJ, Shen J. Current challenges in cardiac rehabilitation: strategies to overcome social factors and attendance barriers. Expert Rev Cardiovasc Ther. 2020 Nov 1;18(11):777–89.

7. Karsten M, Gardenghi G, Arruda ACT, Catai AM, Vieira AM, Stein C, et al. ASSOBRAFIR clinical practice guidelines in cardiovascular physical therapy: Exercise-based interventions in outpatient rehabilitation programs for heart failure. Braz J Phys Ther. 2025 Nov;29(6):101260.

8. Carvalho T de, Milani M, Ferraz AS, Silveira AD da, Herdy AH, Hossri CAC, et al. Diretriz Brasileira de Reabilitação Cardiovascular – 2020. Arq Bras Cardiol. 2020 May 22;114(5):943–87.

9. Yang Z, Zheng X, Hu N, Zhang F, Wang A. “Challenges to Normalcy”- Perceived Barriers to Adherence to Home-Based Cardiac Rehabilitation Exercise in Patients with Chronic Heart Failure. Patient Prefer Adherence. 2023 Dec;Volume 17:3515–24.

10. Fraser MJ, Leslie SJ, Gorely T, Foster E, Walters R. Barriers and facilitators to participating in cardiac rehabilitation and physical activity: A cross-sectional survey. World J Cardiol. 2022 Feb 26;14(2):83–95.

11. da Cruz MMA, Ricci-Vitor AL, Borges GLB, da Silva PF, Turri-Silva N, Takahashi C, et al. A Randomized, Controlled, Crossover Trial of Virtual Reality in Maintenance Cardiovascular Rehabilitation in a Low-Resource Setting: Impact on Adherence, Motivation, and Engagement. Phys Ther. 2021 May 4;101(5).

12. Touloudi E, Hassandra M, Galanis E, Pinnas G, Krommidas C, Goudas M, et al. Effectiveness and acceptance of virtual reality vs. traditional exercise in obese adults: a pilot randomized trial. Front Sports Act Living. 2025 Mar 19;7.

13. Bond S, Laddu DR, Ozemek C, Lavie CJ, Arena R. Exergaming and Virtual Reality for Health: Implications for Cardiac Rehabilitation. Curr Probl Cardiol. 2021 Mar;46(3):100472.

14. Gim G, Bae HK, Kang SA. The Effect of Self-Determination and Quality of VR-Based Education in the Metaverse on Learner Satisfaction. In 2023. p. 41–54.

15. Vermeir JF, White MJ, Johnson D, Crombez G, Van Ryckeghem DML. The Effects of Gamification on Computerized Cognitive Training: Systematic Review and Meta-Analysis. JMIR Serious Games. 2020 Aug 10;8(3):e18644.

16. el Mathari S, Hoekman A, Kharbanda RK, Sadeghi AH, de Lind van Wijngaarden R, Götte M, et al. Virtual Reality for Pain and Anxiety Management in Cardiac Surgery and Interventional Cardiology. JACC: Advances. 2024 Feb;3(2):100814.

17. Costa A dos S, Barbosa CB, Guizilini S, Santos VR dos, Miura CR, Oliveira MT de, et al. Virtual Reality and Physical Activity in Patients with Heart Failure: Technology Validation and User Satisfaction – Pilot Study. International Journal of Cardiovascular Sciences. 2025 Feb 19;38.

18. Kendzierski D, DeCarlo KJ. Physical Activity Enjoyment Scale: Two Validation Studies. J Sport Exerc Psychol. 1991 Mar;13(1):50–64.

19. Alves ED, Panissa VLG, Barros BJ, Franchini E, Takito MY. Translation, adaptation, and reproducibility of the Physical Activity Enjoyment Scale (PACES) and Feeling Scale to Brazilian Portuguese. Sport Sci Health. 2019 Aug 12;15(2):329–36.

20. Brooke J. SUS - A quick and dirty usability scale. 1995.

21. Lourenço DF, Valentim EC, Lopes MHB de M. Translation and Cross-Cultural Adaptation of the System Usability Scale to Brazilian Portuguese. Aquichan. 2022 May 13;22(2):1–16.

22. Forestieri P, Guizilini S, Peres M, Bublitz C, Bolzan DW, Rocco IS, et al. A Cycle Ergometer Exercise Program Improves Exercise Capacity and Inspiratory Muscle Function in Hospitalized Patients Awaiting Heart Transplantation: a Pilot. Braz J Cardiovasc Surg. 2016;

23. Carvalho T de, Freitas OGA de, Chalela WA, Hossri CAC, Milani M, Buglia S, et al. Diretriz Brasileira de Ergometria em População Adulta – 2024. Arq Bras Cardiol. 2024;121(3).

24. Botega NJ, Bio MR, Zomignani MA, Garcia Jr C, Pereira WAB. Transtornos do humor em enfermaria de clínica médica e validação de escala de medida (HAD) de ansiedade e depressão. Rev Saude Publica. 1995 Oct;29(5):359–63.

25. Zigmond AS, Snaith RP. The Hospital Anxiety and Depression Scale. Acta Psychiatr Scand. 1983 Jun 23;67(6):361–70.

26. Castellari C, Barbosa CB. Impact of noninvasive ventilation associated with cycle-ergometer training on exercise tolerance in hospitalized patients with heart failure: a pilot study. . [São Paulo]: Universidade Federal de São Paulo; 2019.

