## Supplemental Table 1 for "REVIVE-HF: Rehabilitation with Immersive Virtual Reality and Exercise in Hospitalized Patients with Heart Failure - A Randomized Controlled Trial Protocol"

Supplementary Table S1. SPIRIT 2025 Structured Summary

| Item | Description |
|---|---|
| Primary Registry and Trial Identifying Number {4} | Brazilian Clinical Trials Registry (ReBEC): RBR-4hrmkzz. https://ensaiosclinicos.gov.br/rg/RBR-4hrmkzz |
| Secondary Identifying Numbers | Ethics approval (CAAE): 54896621.9.0000.5505 |
| Source(s) of Monetary or Material Support | None. Institutional support from Federal University of São Paulo (UNIFESP). |
| Primary Sponsor and contact information {3b} | Name: Federal University of São Paulo (UNIFESP)<br>Contact:<br>Address: Pedro de Toledo Street, 669 - Vila Clementino, São Paulo, SP, Brazil<br>Phone: +55 11 5576-4430 |
| Role of sponsor and funder {3c} | UNIFESP is the responsible institution but has no role in the study design; collection, management, analysis, and interpretation of data; writing of the report; or the decision to submit the report for publication. There is no external sponsor funding this research. |
| Contact for Public Queries | Name: Ariele dos Santos Costa<br>Affiliation: Federal University of São Paulo (UNIFESP)<br>Address: Pedro de Toledo Street, 669 - Vila Clementino, São Paulo, SP, Brazil, CEP 04039-032<br><br>Phone: +55 11 94318-1334 |
| Contact for Scientific Queries | Name: Ariele dos Santos Costa, MSc<br>Position: Principal Investigator<br>Affiliation: Federal University of São Paulo (UNIFESP)<br><br>Phone: +55 11 94318-1334 |

| | |
|---|---|
| | Senior Supervisor:<br>Rita Simone Lopes Moreira, PhD<br> |
| Public Title | Virtual Reality Exercise in Hospitalized Heart Failure Patients |
| Scientific title | REVIVE-HF: Rehabilitation with Immersive Virtual Reality and Exercise in Hospitalized Patients with Heart Failure – A Randomized Controlled Trial Protocol |
| Countries of Recruitment | Brazil |
| Health Condition(s) or Problem(s) Studied | Heart failure (HF) |
| Intervention(s) | **Intervention: Immersive Virtual Reality + Exercise**<br>● Single supervised cycle-ergometer aerobic exercise session<br>● Duration: Up to 20 minutes (five 3-minute pedaling blocks with 1-minute rest intervals)<br>● Technology: VZfit application on Meta Quest 2 headset<br>● Virtual environment: Scenic outdoor routes synchronized with pedaling<br>● Audio: Ambient environmental sounds (no music)<br>● Intensity: Self-selected cadence, RPE-guided (target 3-4 on Modified Borg 0-10 scale)<br>● No external resistance applied<br>● Session terminated if RPE ≥5 or hemodynamic instability<br>**Comparator: Standard Exercise (Control)**<br>● Identical cycle-ergometer protocol without VR<br>● Same duration, intensity monitoring, and safety criteria<br>● Standard hospital environment |

| | |
|---|---|
| Key Inclusion and Exclusion Criteria | ● Ages eligible for study: ≥18 years<br>● Sexes eligible: All<br>● Accepts healthy volunteers: No<br>**Inclusion criteria**<br>● HF with left ventricular ejection fraction (LVEF) ≤ 50% confirmed by echocardiography.<br>● Hemodynamic stability for the previous 48 hours<br>● New York Heart Association (NYHA) Functional Class I-IV (all functional classes included)<br>**Non-inclusion criteria**<br>● Arrhythmias that contraindicate physical exertion.<br>● Unstable angina.<br>● Acute myocardial infarction or cardiac surgery in the previous six months.<br>● Advanced or terminal cancer with life expectancy < 3 months<br>● Cancer-related symptoms preventing safe exercise<br>**Exclusion Criteria**<br>● Fever in the previous 48 hours, with leukocytosis and/or elevated C-reactive protein.<br>● Conditions limiting VR use, specifically: significant visual deficit, cognitive impairment, psychiatric or neurological disorders or history of photosensitive epilepsy.<br>● Orthopedic or neurological conditions precluding safe cycle-ergometer use. |
| Study Type | **Study Design:**<br>● Allocation: Randomized<br>● Intervention model: Parallel assignment |

| | |
|---|---|
| | • Intervention model description: Two-arm, parallel-group randomized controlled trial<br>• Masking description: data analysts blinded to group allocation. Participants and intervention therapists cannot be blinded due to nature of VR intervention.<br>• Primary purpose: Treatment<br>• Study Phase: Not applicable (behavioral/device intervention, not a drug trial)<br>• Enrollment: 60 participants |
| Date of First Enrollment (planned) | • First participant enrollment (planned): January 05, 2026<br>• Estimated study completion date: December 05, 2026<br>• Status: Not yet recruiting |
| Sample Size | • Total: 60 participants<br>• By group:<br>○ Intervention (VR + Exercise): 30 participants<br>○ Control (Exercise only): 30 participants<br>• Allocation ratio: 1:1<br>• Sample size justification: Calculated using G*Power v3.1.9.2 for independent samples t-test (two-tailed, α=0.05, power=0.80). Based on pilot data and prior VR exercise studies showing moderate effect sizes (Cohen's d ≈0.65), 30 participants per group provides adequate power to detect clinically meaningful differences in exercise tolerance. Total of 60 accounts for anticipated 10-15% operational losses/dropout. |
| Primary outcome(s) | • Outcome name: Influence of VR on tolerance to |

| | |
|---|---|
| | cycle-ergometer aerobic exercise<br>● Measure: Cumulative pedaling time (in minutes) from start of exercise to termination due to symptoms or predefined safety criteria<br>● Time frame: Measured during single session (within hospitalization period)<br>● Description: Total time the participant actively pedals before session ends due to: (1) completion of planned protocol (20 minutes/5 blocks), (2) participant request to stop, (3) RPE ≥5 (Modified Borg scale), or (4) hemodynamic instability per safety criteria. This objectively quantifies exercise tolerance in the inpatient setting. |
| Key Secondary outcome(s) | **1. Block-level Physiological Responses**<br>● Systolic and diastolic blood pressure (SBP/DBP, mmHg)<br>● Heart rate (HR, bpm)<br>● Respiratory rate (RR, breaths/min)<br>● Peripheral oxygen saturation ($SpO_2$, %)<br>● Rating of perceived exertion (RPE, Modified Borg Scale 0-10)<br>● Time frame: Baseline, minute 2 of each 3-minute block, -minute post-exercise recovery<br>● Analysis: Repeated measures across timepoints, between-group comparison<br>**2. Exercise Enjoyment**<br>● Measure: Physical Activity Enjoyment Scale (PACES) total score<br>● Time frame: Immediately post-session<br>● Description: 18-item validated questionnaire |

| | |
|---|---|
| | assessing enjoyment during physical activity; higher scores indicate greater enjoyment<br>● Analysis: Between-group comparison<br>**3. VR System Usability**<br>● Measure: System Usability Scale (SUS) total score (0-100)<br>● Time frame: Immediately post-session (intervention group only)<br>● Description: 10-item validated questionnaire; scores >68 indicate above-average usability<br>● Analysis: Descriptive statistics, correlation with tolerance outcomes |
| Ethics Review | ● Status: Approved<br>● Ethics Committee:<br>○ Name: Research Ethics Committee of the Federal University of São Paulo (Comitê de Ética em Pesquisa da UNIFESP)<br>○ Country: Brazil<br>○ Approval number (CAAE): 54896621.9.0000.5505<br>○ Approval date: July 20, 2022<br>○ Contact: |
| Individual Trial Participant Data sharing statement | ● Plan to share IPD: Yes<br>● What will be shared:<br>○ De-identified individual participant data underlying published results (primary and secondary outcomes)<br>○ Data dictionary (variable definitions, coding)<br>○ Study protocol (this document)<br>○ Statistical analysis plan |

| | |
|---|---|
| | ○ Informed consent form (template)<br>○ Analytic code (R or SPSS scripts)<br>● When data will be available:<br>○ Beginning 6 months after publication of primary results<br>○ No end date specified; data will remain available indefinitely via repository<br>● Access criteria:<br>○ Data will be available to anyone who wishes to access the data<br>○ For purposes of reanalysis, meta-analysis, or methodological research<br>○ Via open-access repository (Zenodo) with Digital Object Identifier (DOI)<br>○ No data use agreement or proposal submission required<br>○ All data subject to Creative Commons Attribution 4.0 International License<br>● Supporting information:<br>○ Repository: Zenodo (zenodo.org)<br>○ Accession number: [will be generated upon deposition]<br>○ Persistent identifier: DOI [will be assigned]<br>○ Data format: CSV for datasets, PDF for documents<br>○ Documentation: README file with file descriptions and usage notes<br>● Restrictions:<br>○ No personally identifiable information will be shared (full de-identification per GDPR |

| | |
|---|---|
| | standards)<br>○ No raw unprocessed data from devices if it contains identifiable timestamps or serial numbers<br>○ Video/audio recordings (if any) will not be shared to protect participant confidentiality<br>● Contact for data access:<br>Ariele dos Santos Costa |
